# B and T cell responses after a third dose of SARS-CoV-2 vaccine in Kidney Transplant Recipients

**DOI:** 10.1101/2021.08.12.21261966

**Authors:** Eva Schrezenmeier, Hector Rincon-Arevalo, Ana-Luisa Stefanski, Alexander Potekhin, Henriette Staub-Hohenbleicher, Mira Choi, Friederike Bachmann, Vanessa Pross, Charlotte Hammett, Hubert Schrezenmeier, Carolin Ludwig, Bernd Jahrsdörfer, Andreia C. Lino, Kai-Uwe Eckardt, Katja Kotsch, Thomas Dörner, Klemens Budde, Arne Sattler, Fabian Halleck

## Abstract

**Background:** Accumulating evidence suggests that solid organ transplant recipients, as opposed to the general population, show strongly impaired responsiveness towards standard SARS-CoV-2 mRNA-based vaccination, demanding alternative strategies for protection of this vulnerable group.

**Methods:** In line with recent recommendations, a third dose of either heterologous ChAdOx1 (AstraZeneca) or homologous BNT162b2 (BioNTech) was administered to 25 kidney transplant recipients (KTR) without humoral response after 2 doses of BNT162b2, followed by analysis of serological responses and vaccine-specific B- and T-cell immunity.

**Results:** 9/25 (36%) KTR under standard immunosuppressive treatment seroconverted until day 27 after the third vaccination, while one patient developed severe COVID-19 infection immediately after vaccination. Cellular analysis seven days after the third dose showed significantly elevated frequencies of viral spike protein receptor binding domain specific B cells in humoral responders as compared to non-responders. Likewise, portions of spike-reactive CD4^+^ T helper cells were significantly elevated in seroconverting patients. Furthermore, overall frequencies of IL-2^+^, IL-4^+^ and polyfunctional CD4^+^ T cells significantly increased after the third dose, whereas memory/effector differentiation remained unaffected.

**Conclusions:** Our data suggest that a fraction of transplant recipients benefits from triple vaccination, where seroconversion is associated with quantitative and qualitative changes of cellular immunity. At the same time, the study highlights that modified vaccination approaches for immunosuppressed patients still remain an urgent medical need.

**Significance statement:** Protection of solid organ transplant recipients against SARS-Cov-2 by vaccination remains an unmet need given the low immunogenicity of available vaccines in the presence of immunosuppression. Administration of a third dose to 25 kidney transplant recipients (KTR) resulted in seroconversion in 36% of patients, associated with significant quantitative and functional changes within the spike-antigen-specific B-cell- and CD4^+^ T-helper cell compartment. Our data support the need for individual humoral monitoring of immunosuppressed individuals after vaccination as well as continued efforts to adapt vaccination protocols for this at-risk group.

## Introduction

Vaccines against SARS-CoV-2 have proven high effectiveness among at-risk populations such as the elderly ^1^, patients on maintenance hemodialysis ^2^ or individuals suffering from chronic inflammatory diseases under selected immunosuppressive regimens ^3^. In contrast to the latter, kidney transplant recipients (KTR) receiving immunosuppressive medication exhibit strongly reduced humoral as well as impaired B and T cell responses after mRNA-based vaccination ^4-6^. These observations are in line with reports of breakthrough infections with severe disease courses in fully vaccinated solid organ transplant recipients, underlining the urge of adapted vaccination protocols for this at-risk population ^7, 8^. In order to address this clinical challenge, a third vaccination for solid organ transplant recipients has recently become standard of care in France and Israel ^9^. The first case series reported seroconversion in only 33 % of anti-S1 IgG negative patients after a third vaccine dose, regardless of whether a vector- or an mRNA-based vaccine was used ^8^. These findings are in line with recent data from a larger cohort, where seroconversion increased from 40 % to 67% after three doses of mRNA vaccine ^9^. In the current study, we extend these observations by reporting results in 25 KTR receiving a third vaccination dose of either heterologous ChAdOx1 (Vaxzevria) or homologous BNT162b2 (Comirnaty). In addition to monitoring vaccine-induced humoral responses, we provide comprehensive data on quantitative and qualitative changes within the spike-antigen-specific B- and T cell compartments.

## Results

### Vaccination-induced humoral and B-cell immunity

KTR who received a two-dose BioNTech/Pfizer BNT162B2 immunization three weeks apart and who did not show a humoral response (anti-S1 IgG) were revaccinated with a heterologous ChAdOx1 (n=11, 90±7 days after fist vaccine) or homologous BNT162b2 (n=14, 127±1 days after first vaccine) protocol. In addition to the clinical course humoral, B- and T cell responses were evaluated 7±2 days after the second and third vaccination, and humoral responses were additionally investigated 19-27 days after each vaccination. Anti-S1 specific IgG data were available for all patients, whereas specific IgA and neutralization capacity were only examined in samples from 20 individuals.

One patient developed severe COVID-19 (WHO ordinal scale 6) 10 days after the third vaccination (indicated in red, Figure 1 A-C). 3/25 (12%) KTX patients developed anti-S1 IgG 7±2 days after the third dose, whereas 9/25 (36%) seroconverted until 19-27 days and were categorized as responders. Of these, three patients had received BNT162b2 and six ChAdOx1 as third vaccine; however, anti-S1 IgG was highly positive (OD ratio >5) only in three responders (two BNT162b2 and one ChAdOx1) (Figure 1A). Anti-S1 IgA (Figure 1B) only increased 19-27 days after third vaccination, while neutralization capacity was increased 7±2 days and 19-27 days after the third vaccination as compared to day 7 after the second vaccination, respectively (Figure 1C) in 7/20 patients. The patient with the isolated high anti-S1 IgA had high anti-S1 IgA already before vaccination. The SARS-CoV-2 nucleoprotein was negative at all samplings.

**Figure 1:**
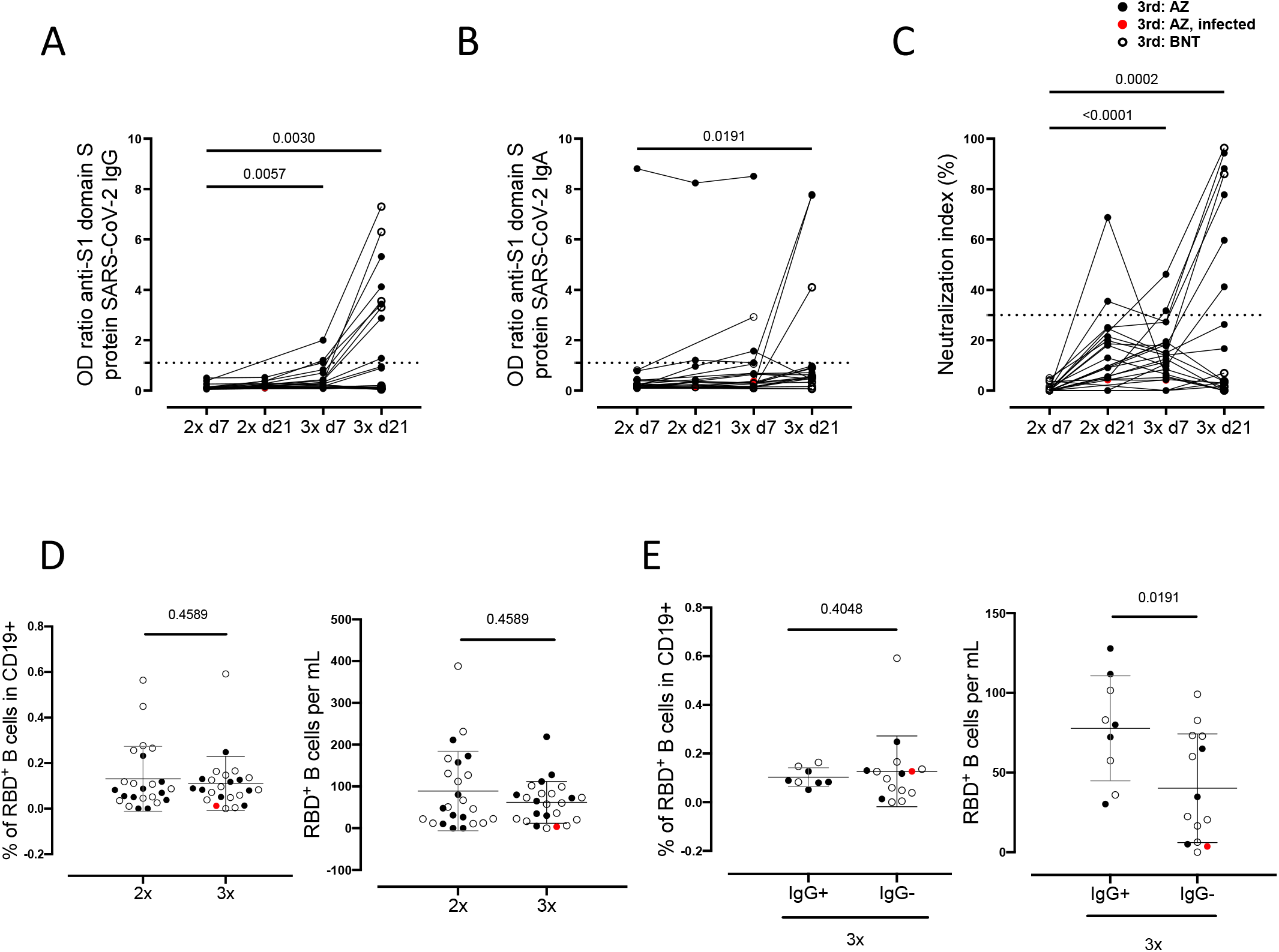
Humoral immune responses and specific B cell immunity after third vaccination in KTR. Humoral vaccine-specific immune responses were assessed by ELISA for anti-spike protein S1 IgG (A), spike protein S1 IgA (B) and virus neutralization by a blocking ELISA (C) at the indicated timepoints in KTR after administration of a third dose of either ChAdOx1 (n=11, black filled dots) or BNT162b2 (n=14, back empty dots). Thresholds defining a positive response are indicated by dotted lines. Relative frequencies (left) and absolute counts (right) of RBD-specific CD19^+^ B cells in all patients (D) as well as in responders IgG+ and non-responders IgG-7±2 days after second or third vaccination with ChAdOx1 or BNT162b2. (A-C) Kruskal-Wallis with Dunn’s post-test. The infected individual is depicted in red. (D, E) Mann-Whitney test.

The relative percentage and absolute number of SARS-CoV-2 spike RBD-specific B cells within the CD19^+^ population did not change between the second and third vaccination (Figure 1D). The relative percentage of SARS-CoV-2 spike RBD-specific CD19 cells did not differ between responders and non-responders (Figure 1E), while absolute numbers of RBD+ B cells were higher among responders compared to non-responders 7±2 days after the third vaccination (Figure 1E).

### Quantitative and qualitative assessment of vaccination-specific CD4^+^ T cell responses

Vaccine-specific CD4^+^ T helper cells were identified within PBMC after stimulation with an overlapping peptide mix encompassing the complete spike glycoprotein based on co-expression of CD137 and CD154, as demonstrated earlier ^6, 10^, and depicted in Supplemental Figure 2. A T cell response was defined as positive when peptide pool stimulated PBMC contained more than twofold higher portions of CD137^+^CD154^+^ T cells as compared to the unstimulated control with at least twenty events. The overall prevalence of vaccinees displaying spike-specific CD4^+^ T cell responses was high (>90%) after the second and third inoculation with no significant changes of antigen-reactive T cell frequencies in cellular responders (Figure 2A). However, individuals who seroconverted after the third vaccination (IgG^+^) were characterized by significantly higher portions of antigen-reactive T cells than humoral non-responders (Figure 2A). Analysis of activation-associated markers revealed a significant drop of proliferating Ki67^+^ and activated PD1^+^ T cells after the third dose (Figure 2B), whereas no changes in specific memory/effector subset composition were noted between the two timepoints (Figure 2C). Importantly, KTR showed significantly higher frequencies of IL-2 and IL-4 secreting as well as polyfunctional IFNγ^+^TNFα^+^IL-2^+^ (triple^+^) T cells after the third dose; this observation did not account for IFNγ or TNFα alone (Figure 2D). Based on the limited sample size, cellular responses were not correlated with the type of the third vaccination.

**Figure 2:**
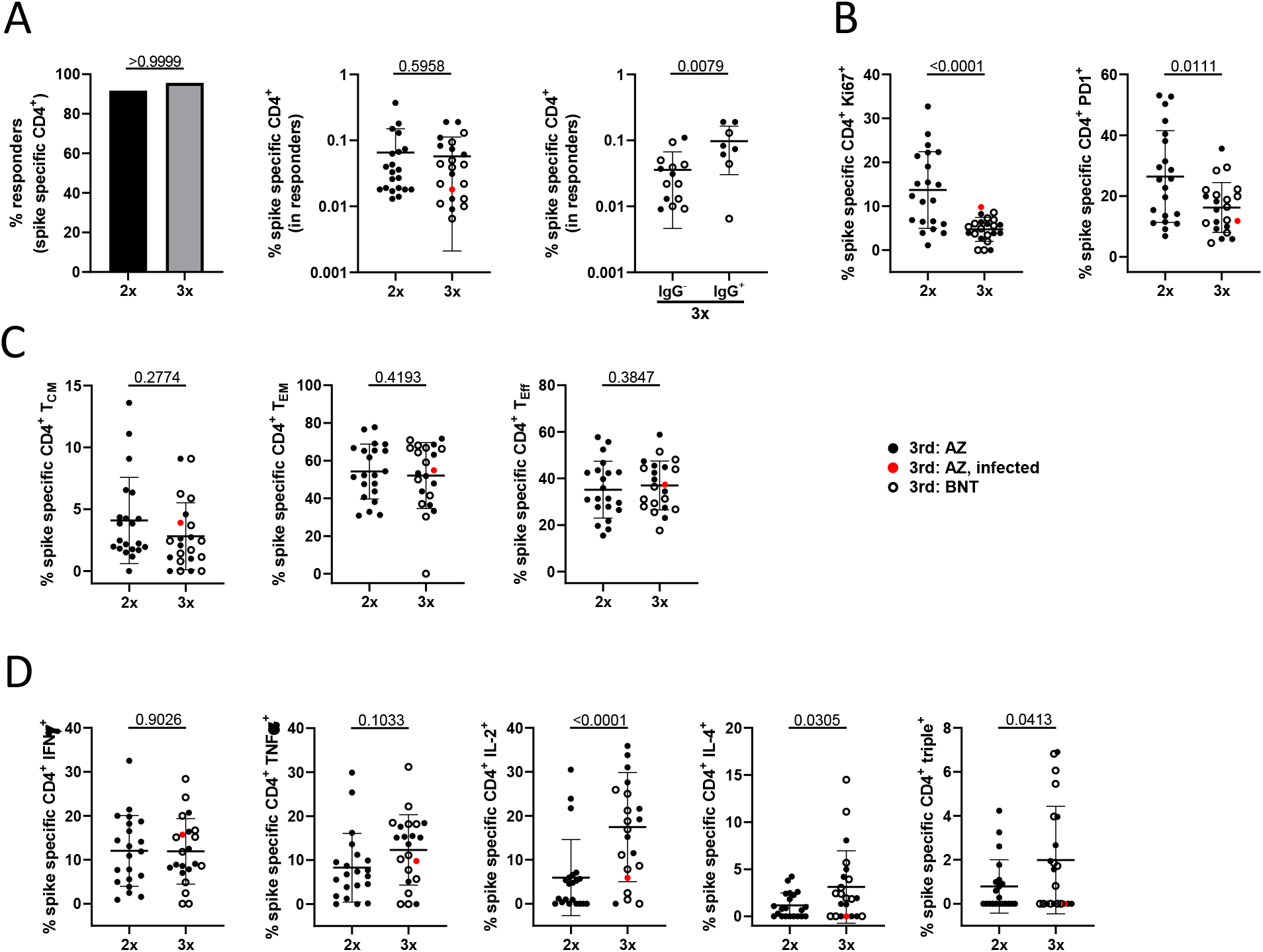
Assessment of T cell reactivity. PBMC of KTX patients (n=24) were stimulated or not with spike peptide mix. Specific CD4^+^ T cells were identified and quantified 7±2 days after the second and third vaccination by FACS according to CD137 and CD154 co-expression. Depicted are (A) the percentage of individuals with a cellular response (left, Fisher’s exact test), frequencies of specific CD4^+^ T cells (middle, paired Wilcoxon test) and frequencies within humoral responders (anti-S1 IgG^+^) and non-responders (anti-S1 IgG^-^) (right, unpaired t test). (B) Frequencies of antigen-specific CD4^+^ T cells expressing Ki67 (left, paired t test) or PD1 (right, paired t test). (C) Memory/effector subset differentiation of spike-reactive CD4^+^ T cells (T_CM_: central memory (left, paired Wilcoxon test), T_EM_: effector memory (middle, paired t test), T_eff_: effector (right, paired t test) T cells). (D) Expression of IFNγ (paired t test), TNFα (paired t test), IL-2 (paird Wilcoxon test) and IL-4 (paired Wilcoxon test in antigen-specific T cells including analysis of IFNγ^+^TNFα^+^IL-2^+^ “triple^+^” polyfunctional (paired Wilcoxon test) cells. Where applicable, graphs show means ± SD.

## Discussion

Investigating the humoral-, B- and T cell response after a third vaccination with either ChAdOx1 or BNT162b2 in previously non-responding KTR we observed an increase in anti-S1 IgG in 9/25 (36%) patients, 4/14 (28%) after homologous and 5/11 (45%) after heterologous vaccination. This observation is in accordance with previously published data of third vaccinations in a heterogeneous group of solid organ transplant recipients ^8, 9^. However, these studies did neither stratify patient responses according to the type of transplant, nor did they quantify vaccine specific cellular T and B cell immunity. There is increasing evidence that neutralizing capacity is a reliable correlate of protection ^11-13^ and that anti-S1 IgG correlates with neutralizing capacity ^14, 15^. Of the 9 responders in our cohort only three developed high titer anti-S1 IgG (OD IgG >5) while one was just above the threshold for positivity. Consistent with lack of protection, one humoral non-responder developed severe COVID-19 after the third vaccination, underlining the clinical importance of adequate antibody titers in KTR, especially with the emergence of viral variants. It has also been reported that antibody titers decline 3 months after vaccination with an mRNA vaccine, so that initial high titers might be necessary for a long term protection ^16^. The absolute number of antigen-specific B cells significantly increased in responders as compared to non-responders after the third dose while it remained largely unchanged compared to the second vaccination. This lack of substantial antigen-specific B and plasmablast induction, as opposed to what has been demonstrated for healthy individuals ^17^, supports the weak additional mobilizing effect of a third vaccination in the majority of KTR and likely explains its overall limited effectiveness.

The exact contribution of vaccine-induced T cell immunity for protection against SARS-CoV-2 infection is still debated, particularly with respect to direct anti-viral effects vs. a role in B cell activation. We recently reported that vaccine-specific CD4^+^ T cells in KTR show broad quantitative and functional limitations ^6^. In line with data on specific B cells presented herein, spike-reactive T cell frequencies were largely unaffected by a third vaccination. In accordance with the fact that an additional boost does not substantially expand the specific T cell pool in KTR, signs of ex vivo activation, as mirrored by high Ki67 or PD1 expression ^6^, were significantly reduced after the third dose as compared to the second dose. For reasons that need to be explored further, humoral responders were characterized by a significant increase of spike-specific T cells, a finding supported by recent work of Sahin et al., demonstrating a strong correlation between specific T cell frequencies and antibody titers post vaccination ^18^. As opposed to their quantities, vaccine-reactive T cells underwent a functional maturation after the third dose with higher portions of IL-2^+^, IL-4^+^ and multifunctional cells. Whereas increased IL-4 secretion could lower the threshold for isotype switching to IgG ^19^, patients might potentially benefit from augmented polyfunctionality being associated with potent SARS-CoV-2 clearance ^20^.

In our cohort, all but one patient received antimetabolite medication. These drugs have been shown to be main contributors to a diminished immune response against SARS-CoV-2 vaccination in solid organ recipients ^21^. One potential strategy to improve responder rates is to reduce immunosuppression and especially antimetabolite treatment in patients with stable allograft function without previous episodes of rejection or pre-existing HLA immunization prior to vaccination. Indeed, data from a large multicenter study suggested that withdrawal of mycophenolate mofetil in KTR formerly receiving tacrolimus-based triple immunosuppression does not impair graft or patient survival ^22^, thereby supporting short-term drug weaning as an option especially in patients that are on corticosteroids as in our cohort.

In conclusion, a third vaccination against SARS-CoV-2 leads to a serological response in a fraction of KTR, while the majority of patients still lacks protective antibody titers. This lack of serological response can be explained by only marginal improvements in cellular protection among antigen-specific B cells and T cells after a third vaccination. Alternative vaccination protocols are urgently needed to protect this at-risk group.

## Materials and Methods

### Study protocol and participants

After standard two-dose BNT162B2 (BioNTech/Pfizer) immunization (21 days apart), 25 KTX patients without anti-spike S1 IgG response received a third vaccination as a standard of care clinical measure under strict monitoring by nephrologists with heterologous ChAdOx1 (n=11, 90±7 days after fist vaccine) or homologous BNT162b2 (n=14, 127±1 days after first vaccine), depending on availability. All participants gave written informed consent for sample collection according to the approval of the ethics committees of the Charité-Universitätsmedizin Berlin (EA2/010/21, EA4/188/20), of the county of Saxony-Anhalt (EA7/21) and the University of Greifswald (BB019/21). Patient demographics are summarized in Table 1. Peripheral blood and serum samples were collected 7±2 days after the second and third vaccination for serological, B and T cell analysis as well as 19-27 days after third vaccination for serology only.

**Table 1:** Patient demographics. Characteristics of kidney transplant (KTX) patients enrolled (n=25)

| Variable |  |
| --- | --- |
| Age (mean yrs ± SD) | 59.7 (13.8) |
| Females (%) | 11 (44.0) |
| Caucasians (%) | 25 (100) |
| **Clinical Parameters** |  |
| Time since Tx (mean yrs ± SD) | 10.4 (8.69) |
| Retransplantation (%) | 5 (20.0) |
| Acute graft rejection (%)<sup>A</sup> | 0 (0) |
| IS medication |  |
| CS+Tac+MMF (%) | 13 (56.0) |
| CS+CyA+MMF (%) | 7 (28.0) |
| mTOR<sub>i</sub>+MMF±CS (%) | 3 (12.0) |
| CyA+mTOR<sub>i</sub> (%) | 1 (4.0) |
| CS+MMF (%) | 1 (4.0) |
| Comorbidities |  |
| Hypertension (%) | 23 (92.0) |
| Coronary heart disease (%) | 6 (24.0) |
| History of myocardial infarction (%) | 2 (8.0) |
| Diabetes (%) | 5 (20.0) |
| History of liver disease (%) | 4 (16.0) |
| History of malignancy (%) | 6 (24.0) |

CS – Corticosteroids, Tac – Tacrolimus, CyA – Cyclosporin A, MMF – Mycophenolate Mofetil, mTOR<sub>i</sub> – mammalian target of rapamycin inhibitor. SD – standard deviation. <sup>A</sup> until 19-27 days after third vaccination

### Serological assessment

SARS-CoV-2 S1 domain specific IgG and IgA was determined by ELISA (Euroimmun). Previous or current SARS-CoV-2 infection was excluded for all but one patient based on medical history in combination with negativity in a SARS-CoV-2 nucleoprotein specific ELISA (Euroimmun). Samples were considered positive with OD ratios of ≥1.1 as per manufacturer’s guidelines. An OD ratio value was determined by calculating the ratio of the OD of the respective test sample over the OD of the internal calibrator provided with the ELISA kit. Virus neutralization capacity of sera was analyzed using a surrogate SARS-CoV-2 neutralization test (GenScript) with more than 30 % being defined as a positive response as described previously ^14, 23^.

### Characterization of antigen-specific B and T cells

All experiments have been performed as previously described ^6, 10, 17^. In brief, peripheral blood mononuclear cells (PBMCs) were isolated by density gradient centrifugation using Ficoll-Paque PLUS (GE Healthcare Bio-Sciences, Chicago, IL, USA).

B cells were detected within PBMC by flow cytometry and gated as CD19^+^CD3^-^CD14^-^ among single live lymphocytes (gating shown in supplementary Figure 1). Antigen-specific B cells were identified by double staining with recombinant purified RBD (DAGC149, Creative Diagnostics, New York, USA) conjugated to AF647 or AF488, respectively.

For analysis of vaccine-specific T cells, 3-5×10^6^ PBMC were stimulated for 16 h with overlapping 15-mers encompassing the complete SARS-CoV-2 spike protein (1 ug/ml per peptide; JPT, Berlin, Germany). Specific CD4^+^ T helper cells were identified based on CD137 and CD154 coexpression as depicted in Supplemental Figure 2. For detection of surface molecules, antibodies against CD3 (SK7, Biolegend, Carlsbad, CA, USA), CD4 (SK3, Becton Dickinson, Franklin Lakes, NJ, USA), CD8 (SK1, Ebioscience, San Diego, CA, USA), CD45RO (UCHL1, BioLegend), CD62L (DREG-56, BioLegend) and PD1 (EH12.1, Becton Dickinson) were used. Unwanted cells were excluded via a “dump” channel containing CD14^+^ (M5E2, BioLegend), CD19^+^ (HIB19, BioLegend), and dead cells (fixable live/dead, BioLegend). After stimulation, cells were fixed in FACS Lysing Solution (Becton Dickinson), permeabilized in FACS Perm II Solution (Becton Dickinson) and intracellularly stained with anti-CD154 (24-31, BioLegend), anti-CD137 (4B4-1, BioLegend), anti-TNF-α (MAb11, BioLegend), anti-IFN-γ (4SB3, Ebioscience), anti-IL-2 (MQ1-17H12, BioLegend), anti-Ki67 (B56, Becton Dickinson), and anti-IL-4 (MP4-25D2, BioLegend). All flow cytometric analyses were performed using a BD FACS Fortessa X20 (BD Biosciences, Franklin Lakes, NJ, USA).

### FACS data analysis and statistics

Flow cytometric data analysis was conducted with FlowJo 10 (Becton Dickinson). The gating strategies for analysis of antigen-specific B- and T cells are illustrated in Supplementary Figure 1 and 2. Co-expression of cytokines was analyzed via Boolean gating. Statistical examination and composition of ELISA and FACS data derived graphs were performed using GraphPad Prism 8 (GraphPad, La Jolla, CA, USA). Parameter distribution was assessed using the Kolmogorov-Smirnov test. Depending on normal distribution or not, a t test or Mann-Whitney test was used for two-group comparisons; for multiple comparisons, a two-way ANOVA with Šidák’s post-test or Kruskal-Wallis test with Dunn’s post-test were chosen. For analysis of contingency tables, Fisher’s exact test was applied.

## Data Availability

All original data are available upon request.

## Acknowledgments

The authors are grateful to Dr. Michael Moesenthin, Dr. Peter Bartsch (both Dialysezentrum Burg), Dr. Ralf Kühn and Dr. Dennis Heutling (both Dialyse Tangermünde) for patient recruitment as well as Dr. Petra Glander and Pia Hambach for biobanking of samples.

## Funding

ES was funded by the Federal Ministry of Education and Research (BMBF) grant (BCOVIT, 01KI20161) and is enrolled in the Charité Clinician Scientist Program funded by the Charité–Universitätsmedizin Berlin and the Berlin Institute of Health. ALS is funded by a scholarship of the German Society of Rheumatology. KK received funding by the Sonnenfeldstiftung Berlin, Germany. TD is grant holder of the Deutsche Forschungsgemeinschaft (Do491/7-5, 10-2, 11-1, Transregio 130 TP24). KK was supported by grants from the Deutsche Forschungsgemeinschaft (KO-2270/71, KO-2270/4-1). AS, KK and FH received project funding from Chiesi GmbH. HRA holds a scholarship of the COLCIENCIAS scholarship No. 727, 2015. HS receives funding by the Ministry for Science, Research and Arts of Baden-Württemberg, Germany and the European Commission (HORIZON2020 Project SUPPORT-E, 101015756).

## Author contribution

ES, AS and FH designed the study and wrote the manuscript.

HRA, ALS, ES and AS performed the experiments (B and T cell analysis)

ES, AP, HSH, FB, KB, CH and MC recruited patients.

HS and BJ were responsible for serological studies.

ES, KB, KK, TD, KUE and FH supervised the work and provided funding.

All authors read and approved the manuscript.

---

**Supplemental Figure 2.**
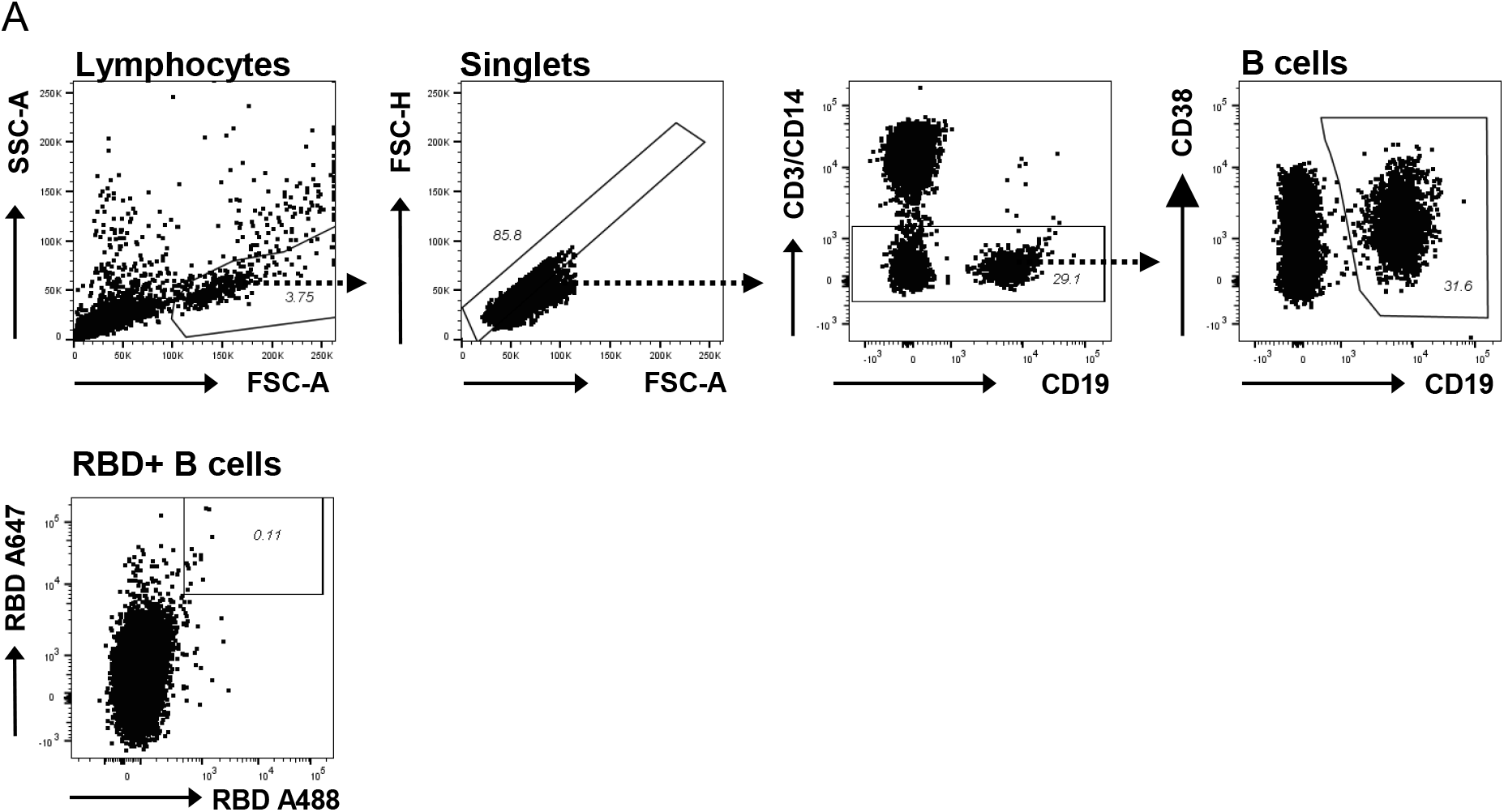
Schrezenmeier, Rincon-Arevalo, *et al* **Detection of SARS-CoV-2 vaccine specific B cells**. B cells in PBMCs were detect by flow cytometry. Antigen-specific B cells were identified by double staining with recombinant purified RBD (DAGC149, Creative Diagnostics, New York, USA) conjugated to AF647 or AF488, respectively.

**Supplemental Figure 2.**
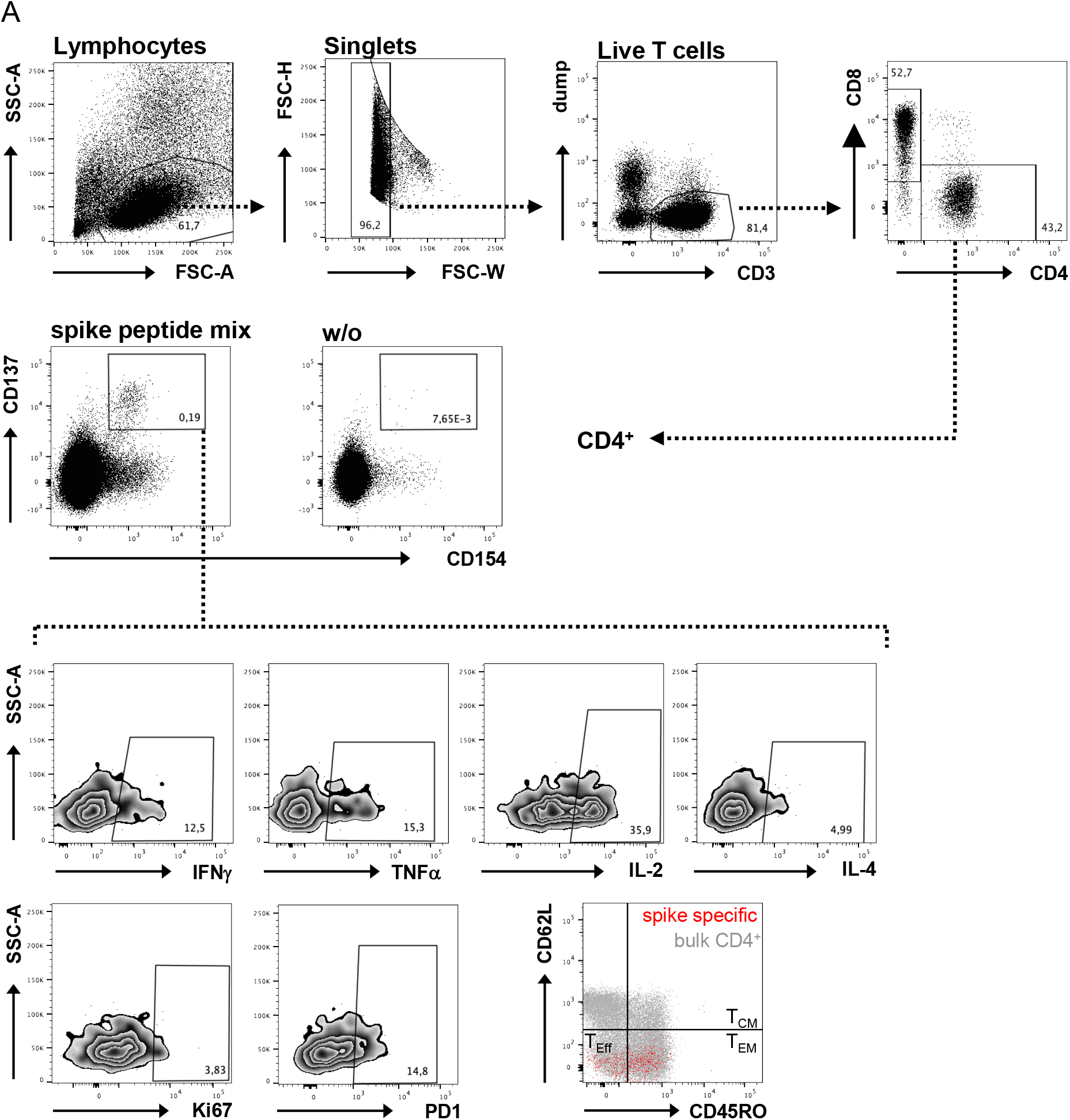
Schrezenmeier, Rincon-Arevalo, *et al* **Detection of SARS-CoV-2 vaccine specific T helper cells**. PBMC were stimulated or not with SARS-CoV-2 spike overlapping peptide mix for 16 h. Antigen-specific live single CD14^-^CD19^-^CD3^+^ (“dump” negative) specific CD4^+^ Th cells were detected by FACS according to co-expression of CD137 and CD154. Specific cells were subsequently analyzed for expression of IFNγ, TNFα, IL-2 and/or IL-4, characterized for expression of the activation-related markers Ki67 and PD1, or for their memory phenotype based on CD45RO and CD62L expression (T_CM_: central memory-, T_EM_: effector memory, T_eff_: effector-T cells). Gates for cytokines or activation-induced molecules were set according to the respective unstimulated or unstained controls, respectively.

## Notes

### Competing Interest Statement

The authors have declared no competing interest.

### Author Declarations

All participants gave written informed consent for sample collection according to the approval of the ethics committees of the Charite Universitaetsmedizin Berlin (EA2/010/21, EA4/188/20), of the county of Saxony-Anhalt (EA7/21) and the University of Greifswald (BB019/21).

